# ETHICAL CONSIDERATIONS FOR CLINICAL CARE ON SHORT TERM MEDICAL MISSIONS IN LOW- AND MIDDLE-INCOME COUNTRIES: A SCOPING REVIEW

**DOI:** 10.1101/2024.02.01.24302164

**Authors:** M. Smith, R. Graham, C. Vance, W. Coburn, D. Richards, J. Whitfield

## Abstract

**Intro:** Despite short term medical missions (STMMs) to low- and middle-income countries (LMIC) becoming increasingly popular, ethical considerations for the provision of clinical care on STMMs are poorly defined. Clinicians are often unprepared to adapt care and ethical precepts to resource limited environments. There may be discord in interpretation of ethical principles between visiting providers and hosts. Clinical care provision has direct impact on the health of patients and communities and there is a need for guidelines regarding ethical clinical care.

**Methods:** Scoping review of the literature published from 2001-2021 restricted to English language identified 3072 records discussing ethical considerations of provision of clinical care on STMMs. Records were screened by title, abstract and finally full text by independent reviewers resulting in 40 records for inclusion.

**Results:** Thirteen themes were identified as important considerations for provision of ethical clinical care on STMMs. These themes included: collaboration/longitudinal relationship, education, lack of follow up, cultural barriers, needs assessment/goal setting, capacity building, outcome evaluation, pre-departure preparation, scope of practice, resources allocation, detriment to local systems, bidirectionality, and formal ethical review. From these themes a list of guidelines is outlined.

**Conclusion:** While ideally clinical care on STMMs would be regulated by formal ethical review boards this is difficult to develop and enforce. Independent STMMs must evaluate their approach to clinical care in LMICs. Care should be given to focusing on collaboration, education, follow up, cultural barriers, and performing a needs assessment/goal setting. These efforts may be guided by the checklist included within.

## BACKGROUND

Medical missions from high income countries (HICs) serve a long-standing role in providing clinical care to patients in low- and middle-income countries (LMICs). In 2012 alone, an estimated 145,185 US physicians provided direct clinical care to patients in LMICs^6^. Clinical environments and resources in LMICs often differ substantially from those in HICs, requiring healthcare practitioners to adapt their practice to new and often complex circumstances. Providers unaccustomed to substantial resource limitations often rely on ethical training that is also based on Western cultural norms in high-resource settings. Even those with the most noble of intentions are limited by the complex interplay of resource limitations, altruism, culture, language, goals, and expectations encountered in such situations. Moreover, the organization-whether private, academic, religious or some combination thereof-sponsoring the medical mission also seeks to fulfill its own goals and expectations, limiting the agency of those they are seeking to serve.

In contrast, application of ethics to medical research in resource-limited settings is a more robustly defined entity. International medical research protocols mandate that any research plan is reviewed by committees with experience in the nuances of medical research in resource-limited settings. As the research commences, there is continuous supervision to ensure adherence to ethical principles for the study’s duration. Unfortunately, similar entities monitoring the ethical provision of clinical care are rare, and this lack of ethical oversight raises concerns for both detected and undetected harm. This is particularly prevalent in short term medical missions (STMMs). Medical mission entities responsible for direct clinical care thus ought to adhere to certain guidelines to ensure provision of ethical care. As such, we performed a scoping review of the literature for ethical considerations pertaining to the provision of clinical care in an emergency medical setting. We then summarized the findings and created a checklist for STMM participants in these settings to help guide ethical care.

## METHODS

A review of the literature published between 2001 and 2021 and restricted to English language was completed to identify articles discussing the ethical considerations of provision of clinical care on STMMs in LMICs. The research did not require ethics review due to lack of human participants. No protocol was filed. Preferred Reporting Items for Systematic Reviews and Meta-Analyses (PRISMA) guidelines were followed throughout. The search was based on elements of emergency treatment and medical missions and included editorials and commentary (APPENDIX 1). The search was completed on 5/6/2021 on EMBASE, PUBMED, Web of Science, and CABI databases. The initial search resulted in 3,107 titles. After removal of duplicates 3,072 titles were available for review. Results pertaining to disaster relief or long-term volunteerism were excluded as the ethical considerations and challenges posed by these mission types are vastly different from those of short-term medical missions and thus outside the scope of this paper. These were divided among five reviewers for inclusion by title. Following this initial review 438 titles remained. These were divided among reviewers and screened by abstract for full text review eligibility. This resulted in 114 articles for full text review. Each full text was reviewed by two independent reviewers. Conflicts were resolved by an independent third reviewer, netting 40 articles that met inclusion. Articles were summarized by reviewers and thematically analyzed.

## RESULTS

The search was conducted on May 6, 2021 resulting in 3,107 records for review. After removal of duplicates and screening of title and abstract for eligibility 114 records were identified for full text review. These were reviewed independently by two reviewers with conflicts resolved by a third reviewer. This resulted in 40 records identified for inclusion.

**Figure 1:**
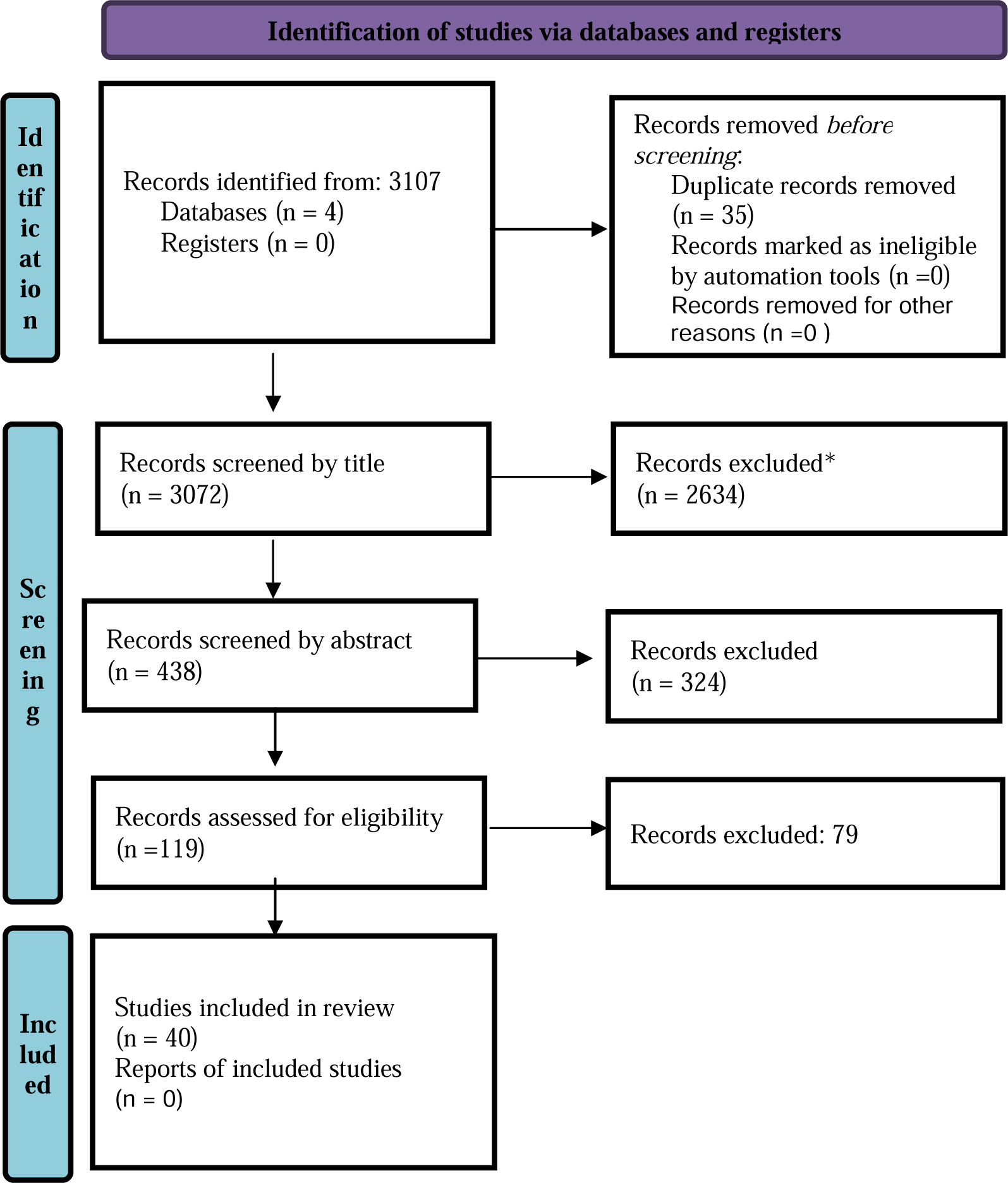
PRISMA flow diagram of screening and review process.

The records were categorized into thirteen themes: collaboration/longitudinal relationship, education, lack of follow up, cultural barriers, needs assessment/goal setting, capacity building, outcome evaluation, pre-departure preparation, scope of practice, resources allocation, detriment to local systems, bidirectionality, and formal ethical review. These themes were defined by all reviewers after the literature review was completed. For the purposes of brevity, the top five themes are discussed within this review. For full list of themes with their identified references and levels of evidence please see APPENDIX 2 and APPENDIX 3 respectively.

**TABLE 1:**
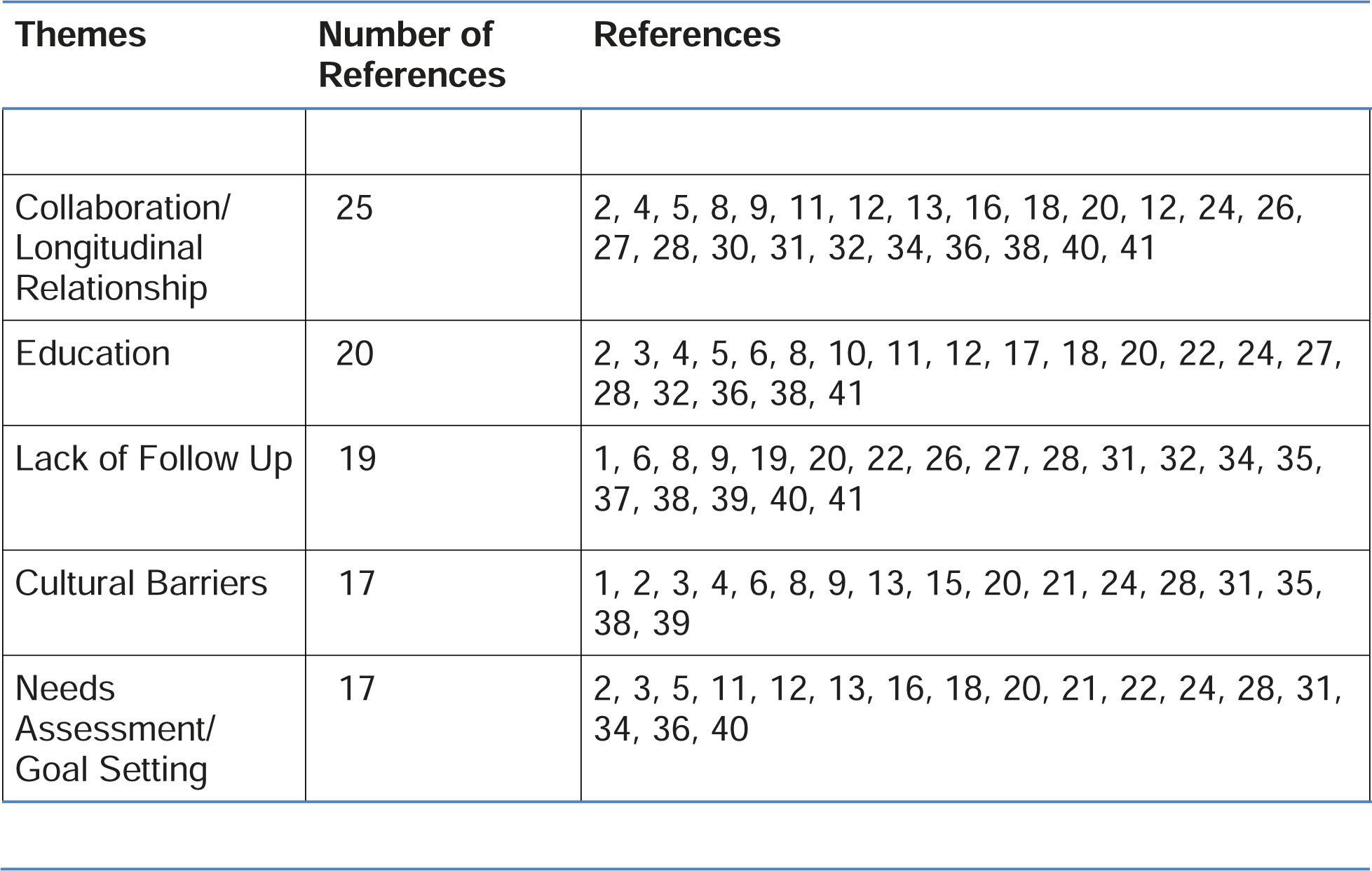
Top Five Identified Themes from the Literature with Associated References.

Other key messages identified included fostering dependence on foreign aid ^1, 32^, STMM influence on structural violence and exacerbating disparities ^9, 13, 15^, issues of informed consent ^9, 29, 31^, solicitation of aid from LMIC’s^11^, review of community perceptions of aid ^26, 39^, improving access to care ^13, 26^, concerns of standard of care^27^, technology mismatch ^38, 40^, cultural differences in ethical frameworks^9^, site selection equity^12^, national incentives for conduct^30^, procedural justice^37^, competitive humanitarianism^38^, and post departure planning^22^.

Records identified for inclusion were sorted by level of evidence based on the following schema: included in level one evidence were Meta analyses, systematic reviews, and randomized controlled trials; level two cohort studies; level three case control studies; level 4 case report or case series; and level 5 narrative reviews, expert opinion, and editorials. The number of records of each level of evidence in support of each category is indicated in TABLE 2.

**TABLE 2:** Levels of Evidence in Support of Top Five Identified Themes.

| Themes | References | #<br>Level<br>1 | #<br>Level<br>2 | #<br>Level<br>3 | #<br>Level<br>4 | #<br>Level<br>5 |
| --- | --- | --- | --- | --- | --- | --- |
| Collaboration/<br>Longitudinal<br>Relationship | 2, 4, 5, 8, 9, 11, 12, 13, 16, 18, 20, 12, 24, 26, 27, 28, 30, 31, 32, 34, 36, 38, 40, 41 | 1 |  |  | 6 | 18 |
| Education | 2, 3, 4, 5, 6, 8, 10, 11, 12, 17, 18, 20, 22, 24, 27, 28, 32, 36, 38, 41 | 1 |  |  | 7 | 11 |
| Lack of Follow<br>Up | 1, 6, 8, 9, 19, 20, 22, 26, 27, 28, 31, 32, 34, 35, 37, 38, 39, 40, 41 | 1 |  |  | 6 | 12 |
| Cultural<br>Barriers | 1, 2, 3, 4, 6, 8, 9, 13, 15, 20, 21, 24, 28, 31, 35, 38, 39 |  |  |  | 9 | 8 |
| Needs<br>Assessment/<br>Goal Setting | 2, 3, 5, 11, 12, 13, 16, 18, 20, 21, 22, 24, 28, 31, 34, 36, 40 |  |  |  | 3 | 14 |

## DISCUSSION

### Collaboration/Longitudinal Relationships

Collaboration and longitudinal relationships was the most frequently identified theme within the literature. Discussion of collaboration and longitudinal relationships encompassed three main components: collaboration with the host country and local community, with other NGO’s or STMMs, and with local health care providers. Strong partnerships with the host country and local community help ensure local needs are met ^2, 11, 20, 27, 30^. This also empowers community members, enhances buy-in, and supports sustainability. Collaboration with the host country can establish assurances that legal conditions are met and potentially result in capacity-building through governmental relationships. Collaboration with other NGOs or STMMs reduces duplicate efforts, allows for coordination and sharing of resources and successes, and offers improved ability to provide follow-up care. Finally, collaboration with local healthcare providers ensures a bidirectional flow of information which results in transfer of skills and knowledge and ensures that culturally competent care is carried out. The literature also suggested that longitudinal relationships improve transfer of knowledge, understanding of local needs, follow-up, outcome evaluation, and capacity-building.

### Education

Education was discussed in two capacities throughout the literature: education of healthcare workers and of community members. Bidirectional teaching between host and visiting healthcare workers was repeatedly emphasized as central to successful missions. As stated by Bermúdez (2004), “education is more than teaching techniques, it is being humble and learning from one another.”^5^ Bidirectional flow of information builds rapport, allows for culturally competent care, and results in capacity-building. A review by Bae et al. (2020) of perspectives of healthcare workers in LMIC’s with experience in STMMS found that subspecialty education is the most desired attribute of STMMS by LMIC providers.^3^ Education of local providers can portend sustainable changes and better patient outcomes. The educational benefit to the volunteers from HIC’s is repeatedly mentioned; particularly the value of learning opportunities provided for students and benefits of seeing conditions not prevalent in HIC’s. Such opportunities must be reviewed to ensure that the volunteer’s educational benefit does not overshadow the needs and benefits for the community or result in harm. For example, students learning procedures outside of their scope or skill set. Other suggestions include the development of international internships or residencies for LMIC physicians in HICs. These opportunities could provide education and allow for the translation of curricula and training to the LMIC. When considering educational benefits for local communities one should aim to build health literacy programs. Considerations should include making educational messages available in the local language and dialect, take into account locally available resources, and be culturally sensitive.

### Follow Up

Lack of follow-up after a STMM was frequently cited as leading to poor outcomes for patients. As recognized by McCurry and Aldulaimi (2018), the value of interventions performed during a STMM are called into question when a patient is unable to receive adequate continuity of care^22^. This includes the inability to refill medications, monitor health conditions, and resolve complications. The latter was noted as a particular concern for surgical procedures with which local providers may be unfamiliar.

Furthermore, lack of follow-up may lead to use of expired supplies or defective equipment left behind by STMMs.^19^ In addition, a survey of local providers by Nouvet et al. (2018), it was noted that care provided to a patient on a STMM is not always communicated to the local team and is therefore difficult to integrate into ongoing care.^26^ Finally, there is a lack of outcomes evaluations of STMMs, so their impact is not well understood.

Suggestions for improving follow-up lie heavily on close partnerships between communities and local providers. Local providers should oversee all care provided on STMM so that monitoring requirements, new findings and treatments, or procedural complications are known. This can further aid in appropriate referrals being made for continuity of care as local providers will have a deeper understanding of local infrastructure. Roche et al. (2017) noted in their systematic review that “about a third of articles (29 %, 27/92) recommend that [STMMs] consider the feasibility of follow-up care after their departure, with several acknowledging this may require hiring local doctors to help. [STMMs] sometimes encounter patients who require care beyond what the [STMM] can offer; however, only six publications (7 %) recommend that [STMMs] refer such patients to the local healthcare system.”^27^

When conducting pre departure planning, plans for follow-up should be considered. Potential interventions should be evaluated for their need for ongoing care and the capacity of local resources to provide that care. One potential solution is for better follow up structuring and outcome evaluation in STMM’s that returns to one site repeatedly to further build relationships with patients and providers.

### Cultural Barriers and Humility

Much of the literature supported the notion that language barriers, along with lack of cultural humility and understanding of local systems, can result in ineffective patient care due to miscommunication and poor sensitivity.^1, 2,13, 15, 20, 28, 31, 35^ Language barriers in particular are detrimental to patient-provider and provider-provider relationships.^13, 15, 20, 21, 24, 28^ Stone and Olsen (2016) note that “cultural differences may make communication and understanding difficult with respect to expectations, values, and decision making.”^31^ This holds true both in individual treatment and when considering the needs and goals of the host community.^20^ Furthermore, language barriers can perpetuate structural violence^15^ as consent and power structures are called into play. Lack of respect for and knowledge of local cultural norms may also be detrimental to relationships between the provider and the local community. As Wall (2011) notes, some cultures attribute supernatural or religious belief to illnesses. By failing to evaluate patient perceptions, providers forgo opportunities to engage in shared decision-making and find treatments amenable to patient’s belief systems.^35^ Furthermore, misunderstanding of customs may result in offending a patient and degrading a patient-provider relationship.^28^ Customs regarding hospitality can lead to the displacement of local providers or impede criticisms and suggestions for improvement for the STMM.^20^ The resounding suggestion to improve cultural humility is through pre-departure training in basic customs, work conditions, language skills, and medical conditions. This demonstrates a flexibility and willingness for bidirectional learning thereby improving relationships.^3^ Notably, while proponents of pre-departure training in cultural humility agree that professionalism and ethics should follow STMM providers, it remains important to note that many cultures view ethical precepts differently.^9^ As such, STMM providers should be aware of and prepared for ethical dissonance and work to understand what those in the host country value in order to foster collaboration. For example, Coors et al. (2015) found that in choosing patients for heart surgery there was a dissonance in how patients were chosen between host and visiting providers. The Rwandan providers prioritized patients based on socioeconomic status based on their value placed on productivity. This was particularly true as the principle of harm was viewed by hosts as an intervention “outpacing the individual patient’s opportunity for meaningful work… improved health but no improvement in overall economic, career or educational circumstances”^9^. In contrast, visiting providers perception concerning justice was that everyone should have an equal chance at surgery. Coors et al. (2015) also found that the Rwandan program believed they discouraged collaboration and return of visiting teams by revealing these productivity - based standards for selection.

### Needs Assessment

Performing a needs assessment within a community is regarded as critical to establishing goals. As Dearani et al. (2016) suggests, realistic expectations and goals should be set at the beginning of a partnership and focus on the needs of a community and an NGO’s ability to address those needs.^11^ This may define the composition of a visiting team and help determine what is required for success. This includes an evaluation of available resources-for instance, as DeCamp et al. (2014) found -there was already infrastructure available for community programs that was unused.^13^ Needs assessments can aid local capacity-building by focusing on supporting local programs for health promotion already in place. ^22, 24^ Respect for local needs, resources, and health promotion drives may also foster relationships particularly when goals of the volunteers and hosts conflict.^3^ In fact, in Bae’s (2020) study of 102 host country physicians, 80% of survey respondents supported development of a platform on which they could advertise for their needs rather than having generic STMMs.^3^ A needs assessment should include all stakeholders in the conversation including community leaders, health officials and community members who may act as health representatives.^18, 20^

### Summary of findings

In order to provide ethical care within STMMs in LMICs programs should aim to build longitudinal bidirectional educational partnerships with LMICs. We propose that this can be accomplished by attending to the following checklist of considerations.

- Begin with conducting a collaborative needs assessment with the host country stakeholders.

○ Consider identifying a community leader for outreach
- Use needs assessment to guide goal setting together with the host country professionals

○ Consider goals which aim to build local capacity
○ Evaluate goals to ensure that they don’t impede local systems or duplicate local efforts
○ Ensure that goals are focused on needed interventions and are attainable
○ Solicit educational goals from the host country professionals-i.e. specialty education, procedural
○ Identify educational goals for visiting professionals-i.e. gaining knowledge of local illness or beliefs
- Conduct pre departure planning to develop a framework to achieve set goals, including:

○ Inventory of available resources, including those needed for follow-up
○ Scope and level of training of volunteers

▪ Deference to host country professionals as local experts
▪ Host country professionals should lead clinical care, visiting professionals acting in support.
○ Plan for outcome evaluation
○ Components of consent for interventions and provision in local language
○ Educational tools or handouts in local languages
- Conduct pre-departure training to include information on basic customs, work conditions and available resources, basic language skills, and common medical conditions.

○ Ideally developed with locals from the host country
- Arrange for patient follow-up either with local resources or on a planned return trip
- Conduct outcome evaluation, use this to guide future efforts.

○ Ideally STMMs will return to the same community

## LIMITATIONS

Limitations in the development of this checklist include the lack of level one evidence available on this topic. The majority of articles identified for full text review were level four and five evidence with only three articles identified as level one evidence. This is inherently expected due to the complex nature of the topic of ethics and the reason commentary and other level four and five evidence were included in the initial search. Furthermore, the lack of higher-level evidence indicates a need for further research and consensus in this area. The articles identified as level one evidence did support the ideas of the top identified themes of collaboration/longitudinal relationships, education, and lack of follow up as well as themes of bidirectionality, pre-departure planning, and formal ethical review.

## CONCLUSIONS

Ideally, clinical care would adhere to the same rigorous ethical review as research missions. Clinical review boards would likely aid in reducing potential harms and contribute to the success of creating and achieving appropriate mission goals by requiring pre-mission planning and post-mission evaluations. As not all STMMs are associated with larger academic entities, however, this is difficult to arrange and enforce. Additionally, it would require significant financial and personnel resources which may detract from care provision.

In the absence of review boards, it falls to individual groups to ensure provision of ethical care. A review of the literature identified collaboration, education, follow-up, cultural barriers and needs assessment as the most commonly identified components of considerations necessary to provide ethical care in LMICs on STMMs. Discussions within the literature of these themes allowed for identification of a checklist of considerations for provision of ethical care which may be referenced when developing a STMM.

## Data Availability

All data produced in the present study are available upon reasonable request to the authors

## APPENDIX 1: Search Terms

### PubMed

(Emergency Treatments[tiab] OR Emergency Therapy[tiab] OR Emergency Therapies[tiab] OR Emergency care[tiab] OR Emergency treatment[tiab] OR Emergency medicine[tiab] OR “Emergency Treatment”[Mesh] OR “Emergency Medicine”[Mesh] OR “Emergency Medical Services”[Mesh] OR “ambulance” OR “emergency medical technician” OR “emt” OR “paramedic” OR “prehospital” OR “pre hospital”) AND (“Comment” [Publication Type] OR commentary[tw] OR “consensus”[tw] OR “Editorial” [Publication Type] OR editorial[tw] OR editor opinion[tw] OR editorial comment[tw] OR “Consensus Development Conference” [Publication Type] OR “Consensus Development Conference, NIH” [Publication Type] OR consensus development[tw]) AND Medical missions[tiab] OR Medical Mission[tiab] OR humanitarian aid workers[tiab] OR voluntarism[tiab] OR Medical humanitarian missions[tiab] OR Medical humanitarian mission[tiab] OR Medical relief[tiab] OR Volunteerism[tiab] OR “Medical Missions”[Mesh]

### EMBASE

((‘Emergency Treatments’ OR ‘emergency therapy’ OR ‘Emergency therapies’ OR ‘emergency care’ OR ‘emergency treatment’ OR ‘emergency medicine’ OR ‘ambulance’ OR ‘emergency medical technician’ OR ‘emt’ OR ‘paramedic’ OR ‘prehospital’ OR ‘pre hospital’):ab,ti OR ‘Emergency Treatment’/exp OR ‘Emergency Medicine’/exp) AND (‘Editorial’:it OR ‘commentary’:de,ab,ti OR ‘consensus’:de,ab,ti OR ‘editorial’:de,ab,ti OR ‘editor opinion’:de,ab,ti OR ‘editorial comment’:de,ab,ti OR ‘consensus development’:de,ab,ti OR ‘letter’:it) AND (‘Medical missions’:ab,ti OR ‘Medical Mission’:ab,ti OR ‘humanitarian aid workers’:ab,ti OR ‘voluntarism’:ab,ti OR ‘Medical humanitarian missions’:ab,ti OR ‘Medical humanitarian mission’:ab,ti OR ‘Medical relief’:ab,ti OR ‘Volunteerism’:ab,ti OR ‘international cooperation’/exp OR ‘international organization’/exp)

### CABI

title:((‘Emergency Treatments’ OR ‘emergency therapy’ OR ‘Emergency therapies’ OR ‘emergency care’ OR ‘emergency treatment’ OR ‘emergency medicine’ OR ‘ambulance’ OR ‘emergency medical technician’ OR ‘emt’ OR ‘paramedic’ OR ‘prehospital’ OR ‘pre hospital’)) OR ab:((‘Emergency Treatments’ OR ‘emergency therapy’ OR ‘Emergency therapies’ OR ‘emergency care’ OR ‘emergency treatment’ OR ‘emergency medicine’ OR ‘ambulance’ OR ‘emergency medical technician’ OR ‘emt’ OR ‘paramedic’ OR ‘prehospital’ OR ‘pre hospital’))

AND title:(‘commentary’ OR ‘consensus’ OR ‘editorial’ OR ‘editor opinion’ OR ‘editorial comment’ OR ‘consensus development’) OR ab:(‘commentary’ OR ‘consensus’ OR ‘editorial’ OR ‘editor opinion’ OR ‘editorial comment’ OR ‘consensus development’) OR it:(‘commentary’ OR ‘consensus’ OR ‘editorial’ OR ‘editor opinion’ OR ‘editorial comment’ OR ‘consensus development’)

AND title:(‘Medical missions’ OR ‘Medical Mission’ OR ‘humanitarian aid workers’ OR ‘voluntarism’ OR ‘Medical humanitarian missions’ OR ‘Medical humanitarian mission’ OR ‘Medical relief’ OR ‘Volunteerism’) OR ab:(‘Medical missions’ OR ‘Medical Mission’ OR ‘humanitarian aid workers’ OR ‘voluntarism’ OR ‘Medical humanitarian missions’ OR ‘Medical humanitarian mission’ OR ‘Medical relief’ OR ‘Volunteerism’)

### WEB of Science

TS=((‘Emergency Treatments’ OR ‘emergency therapy’ OR ‘Emergency therapies’ OR ‘emergency care’ OR ‘emergency treatment’ OR ‘emergency medicine’ OR ‘ambulance’ OR ‘emergency medical technician’ OR ‘EMT’ OR ‘paramedic’ OR ‘prehospital’ OR ‘pre hospital’))

AND TS=(‘commentary’ OR ‘consensus’ OR ‘editorial’ OR ‘editor opinion’ OR ‘editorial comment’ OR ‘consensus development’)

AND TS=(‘Medical missions’ OR ‘Medical Mission’ OR ‘humanitarian aid workers’ OR ‘voluntarism’ OR ‘Medical humanitarian missions’ OR ‘Medical humanitarian mission’ OR ‘Medical relief’ OR ‘Volunteerism’)

## APPENDIX 2: Identified themes from the literature with associated references

| Themes | Number of References | References |
| --- | --- | --- |
| Collaboration/<br>Longitudinal<br>Relationship | 25 | 2, 4, 5, 8, 9, 11, 12, 13, 16, 18, 20, 12, 24, 26, 27, 28, 30, 31, 32, 34, 36, 38, 40, 41 |
| Education | 20 | 2, 3, 4, 5, 6, 8, 10, 11, 12, 17, 18, 20, 22, 24, 27, 28, 32, 36, 38, 41 |
| Lack of Follow Up | 19 | 1, 6, 8, 9, 19, 20, 22, 26, 27, 28, 31, 32, 34, 35, 37, 38, 39, 40, 41 |
| Cultural Barriers | 17 | 1, 2, 3, 4, 6, 8, 9, 13, 15, 20, 21, 24, 28, 31, 35, 38, 39 |
| Needs<br>Assessment/<br>Goal Setting | 17 | 2, 3, 5, 11, 12, 13, 16, 18, 20, 21, 22, 24, 28, 31, 34, 36, 40 |
| Capacity Building | 16 | 1, 2, 5, 6, 8, 9, 11, 12, 19, 20, 21, 24, 25, 28, 30, 36 |
| Outcome<br>Evaluation | 14 | 2, 6, 11, 12, 13, 16, 20, 24, 28, 30, 31, 34, 37, 40 |
| Pre-Departure<br>Preparation | 14 | 2, 3, 6, 8, 11, 14, 15, 23, 24, 26, 28, 30, 33, 35 |
| Scope of Practice | 15 | 3, 4, 8, 13, 14, 15, 21, 24, 28, 30, 31, 32, 35, 37, 40 |
| Resource<br>Allocation | 10 | 5, 8, 9, 19, 21, 27, 31, 32, 35, 39 |
| Detriment to Local<br>Systems | 8 | 1, 4, 6, 16, 20, 28, 32, 40 |
| Bidirectionality | 5 | 3, 5, 11, 17, 24, 27 |
| Formal Ethical<br>Review | 5 | 12, 16, 20, 27, 36 |

## APPENDIX 3: Levels of Evidence in Support of Identified Themes

| Themes | References | #<br>Level<br>1 | #<br>Level<br>2 | #<br>Level<br>3 | #<br>Level<br>4 | #<br>Level<br>5 |
| --- | --- | --- | --- | --- | --- | --- |
| Collaboration/<br>Longitudinal<br>Relationship | 2, 4, 5, 8, 9, 11, 12, 13, 16, 18, 20, 12, 24, 26, 27, 28, 30, 31, 32, 34, 36, 38, 40, 41 | 1 |  |  | 6 | 18 |
| Education | 2, 3, 4, 5, 6, 8, 10, 11, 12, 17, 18, 20, 22, 24, 27, 28, 32, 36, 38, 41 | 1 |  |  | 7 | 11 |
| Lack of Follow<br>Up | 1, 6, 8, 9, 19, 20, 22, 26, 27, 28, 31, 32, 34, 35, 37, 38, 39, 40, 41 | 1 |  |  | 6 | 12 |
| Cultural Barriers | 1, 2, 3, 4, 6, 8, 9, 13, 15, 20, 21, 24, 28, 31, 35, 38, 39 |  |  |  | 9 | 8 |
| Needs<br>Assessment/<br>Goal Setting | 2, 3, 5, 11, 12, 13, 16, 18, 20, 21, 22, 24, 28, 31, 34, 36, 40 |  |  |  | 3 | 14 |
| Capacity<br>Building | 1, 2, 5, 6, 8, 9, 11, 12, 19, 20, 21, 24, 25, 28, 30, 36 |  |  |  | 5 | 11 |
| Outcome<br>Evaluation | 2, 6, 11, 12, 13, 16, 20, 24, 28, 30, 31, 34, 37, 40 |  |  |  | 3 | 11 |
| Pre-departure<br>Preparation | 2, 3, 6, 8, 11, 14, 15, 23, 24, 26, 28, 30, 33, 35 | 2 |  |  | 5 | 7 |
| Scope of<br>Practice | 3, 4, 8, 13, 14, 15, 21, 24, 28, 30, 31, 32, 35, 37, 40 |  |  |  | 6 | 9 |
| Resource<br>Allocation | 5, 8, 9, 19, 21, 27, 31, 32, 35, 39 |  |  |  |  |  |
| Detriment to<br>Local Systems | 1, 4, 6, 16, 20, 28, 32, 40 |  |  |  | 4 | 4 |
| Bidirectionality | 3, 5, 11, 17, 24, 27 | 1 |  |  | 1 | 3 |
| Formal Ethical<br>Review | 12, 16, 20, 27, 36 | 1 |  |  |  | 4 |

